# Predicting county-level opioid overdose mortality in Ohio through multimodal data: A spatiotemporal study using toxicology and EMS indicators, 2020-2024

**DOI:** 10.64898/2026.11.18.26368445

**Authors:** Joanne Kim, John V. Myers, Charles Marks, Brandon Slover, Penn Whitley, Xianhui Chen, Changchang Yin, Ping Zhang, Neena Thomas, Tim Huerta, Eric Dawson, Naleef Fareed, Soledad Fernandez

## Abstract

**Background:** The opioid crisis is the leading driver of overdose mortality in the United States. While overdose deaths have decreased in recent years, the reporting lag in official overdose deaths presents a hurdle in implementing public health interventions to combat the crisis.

**Objective:** To address these data-reporting delays and improve prediction during highly volatile periods, we developed a Bayesian hierarchical spatiotemporal model using real-time drug overdose data sources to predict quarterly overdose deaths across Ohio counties from 2020 to 2024.

**Methods:** We implemented a Bayesian spatiotemporal model using Integrated Nested Laplace Approximation (INLA). Rapid-reporting surveillance metrics, specifically urine drug test (UDT) positivity rates and Emergency Medical Services (EMS) naloxone administration events, were incorporated as fixed covariates. Models were constructed stepwise by first optimizing the spatiotemporal random effect structure and then sequentially adding fixed effects to minimize prediction error. A rolling training window was implemented and evaluated across different training window sizes. Results were assessed overall and stratified by county population size.

**Results:** We found that the optimal predictive model structure consists of a two-year training window, a random walk temporal effect, a structured spatial effect, and fentanyl UDT positivity and EMS naloxone events. Incorporating these UDT and EMS data significantly enhanced predictive performance, with the most pronounced accuracy gains occurring during the unprecedented overdose increase associated with the COVID-19 pandemic in the second quarter of 2020. While predictive performance was highly robust in large, urban counties, predictive capability in smaller counties was limited by low data availability.

**Conclusion:** Our spatiotemporal prediction model successfully bridges the administrative reporting lags and offers near-real-time quarterly overdose prediction. The study also showed that integration of real-time data sources is especially beneficial for capturing sudden, volatile spikes in overdose deaths.

## Introduction

The opioid crisis is a pressing public health challenge in the United States, as opioid-related overdose deaths have contributed to the majority of overdose deaths since the 2000s. Although Ohio opioid overdose deaths have decreased since peaking in 2021-2022, a lag in overdose death reporting complicates efforts to combat the crisis. The Centers for Disease Control (CDC) Wonder database historically updated overdose death data with a six-month lag. The State of Ohio Integrated Behavioral Health Dashboard (IBHD), while inactive since February 2025 (as last verified in July 2026), had reported overdose deaths with a two-month reporting lag [1]. Despite shortened reporting windows, the reporting gap makes it difficult to respond to the crisis in a timely manner. Although the IBHD contains measures such as opioid overdose deaths, naloxone units distributed, and incidents of higher dosage of opioids, it lacks information that directly proxies near-real-time illicit use of opioids in the community. In this paper, we propose a Bayesian spatiotemporal modeling approach to generate up-to-date county-level predictions at quarterly intervals using near-real-time drug positivity and treatment data, and sociodemographic data. Past county-level prediction studies used socioeconomic data such as the American Community Survey (ACS) and drug overdose-related healthcare data [2, 3]. In this study, we incorporated urine drug test (UDT) data in addition to sociodemographic and health department data. Clinicians in general healthcare facilities and substance use treatment and rehabilitation facilities routinely order UDT. After that, drug testing labs such as Millennium Health analyze samples, de-identify, and aggregate UDT data. UDT is thought to serve as a timely proxy measure for drug use in communities; previous research established strong correlations between the percentage of positive UDT results and overdose death at the national and regional levels [4, 5]. We previously demonstrated that UDT assists with opioid overdose death prediction at the county and national level [6, 7]. In one such study, our generalized linear model using fentanyl-related UDT measures exhibited improved performance over an ARIMA model [7]. We observed that higher temporal resolutions, such as monthly or quarterly intervals, exhibit far more pronounced fluctuations in overdose mortality compared to smoothed annual trends. Consequently, we hypothesized that the near-real-time availability of UDT data would be instrumental in capturing these acute, highly volatile shifts in overdose deaths.

Bayesian models that incorporate spatiotemporal random effects with prior distributions can capture underlying variations related to geographic distribution in deaths. This approach has been widely used for studying opioid overdose deaths for public health surveillance to identify risk factors in retrospective studies [8] and predicting future overdose death trends [9]. However, existing Bayesian spatiotemporal forecasting models for opioid overdose mortality have largely been restricted to pre-2020 data or have focused on annual-level predictions [2, 3, 9]. Such annual models are inherently limited in their capacity to detect rapid, short-term fluctuations or sudden shifts in overdose trends. As multiple studies and data sources have reported [10], opioid-related mortality underwent a substantial escalation during the COVID-19 pandemic. Trends from 2020 show heightened volatility, necessitating the development of robust statistical frameworks capable of rapidly adapting to emerging shifts.

This paper aims to pursue two objectives. First, we develop a Bayesian spatiotemporal model to predict overdose deaths across Ohio to overcome reporting gaps and provide near-real-time prediction results. Second, we investigate drug positivity and EMS data in the prediction model to account for overdose death volatility, together with overdose treatment and sociodemographic data. We hypothesized that integrating rapid-reporting surveillance metrics—specifically UDT positivity rates and EMS event data—would enhance model performance during periods of high epidemiological volatility, reflecting a strong association between these proxy measures and fatal opioid overdoses.

## Methods

### Data

#### I. Ethical Consideration

Authorization for data utilization was obtained from The Ohio State University IRB (Study# 2022H0445 approved on 5/2/2023), the Ohio Department of Health Institutional Review Board (The Ohio Department of Health Human Subjects Institutional Review Board *(FWA00001963, IRB00002180)* approved 12/03/2024), and the data were accessed for research purposes on 04/29/2025. The requirement for informed consent was waived by the reviewing institutional review board(s) because the study involved retrospective analysis of existing data. Study data were accessed and used in accordance with applicable institutional data-use requirements and the approved data use agreements. State data obtained from the Ohio Department of Health were accessed and used in accordance with the applicable ODH data-use requirements. The authors did not have access to information that could directly identify individual participants during or after the study.

#### II. Ohio opioid overdose death and other state data

Opioid overdose deaths were obtained from death records provided by the Ohio Department of Health (ODH) from January 2015 through December 2024. The extensive data, dating back to 2015, enabled us to investigate the different lengths of the training period for our model. Eligible mortalities must have Ohio residency and be over 18 years of age at the time of death. We first narrowed the death record data down to drug overdose deaths, defined as a mortality containing an ICD10 code [11] of X40-X44 (accidental poisoning), X60-X64 (intentional poisoning), X85 (homicide by poisoning), or Y10-Y14 (poisoning where the cause of death was unclear) as a primary or secondary cause of death. Within the scope of overdose deaths, opioid involvement is defined as having a code T40.0-T40.4 or T40.6 (poisoning related to narcotics). Deaths were aggregated to the quarterly level by county of residence.

Social Deprivation Index (SDI) measures were sourced from the Graham Center [12]. SDI measures are derived from the 2015-2019 American Community Survey (ACS) to measure socioeconomic disadvantage in a community. The overall SDI measure is expressed as a percentile with 1 being the most advantaged socioeconomically and 100 being the most disadvantaged. We also sourced EMS events with naloxone administration, EMS events with suspected opioid poisoning, naloxone units distributed by Project DAWN, and measures related to medication for Opioid Use Disorder (OUD) and safer prescribing as overdose death treatment effects. The EMS events act as a near-real-time proxy for the total volume of acute opioid overdose deaths [13]. All EMS variables were obtained from the State of Ohio Integrated Behavioral Health Dashboard [1]. IBHD data were reported monthly and were lagged by two months.

#### III. Urine drug test data

Real-time drug use data from 2015 to 2024 are provided by Millennium Health (MH) LLC, an accredited laboratory offering UDT services across the country. The study protocol to use the UDT was approved by the Aspire independent review board, which included a waiver of consent for the use of de-identified data. Specimens for UDT were submitted by health care professionals to Millennium Health as part of a patient care plan between January 2015 and December 2024. Analyses used definitive test results performed by liquid chromatography tandem mass spectrometry for fentanyl (fentanyl or norfentanyl metabolite), cocaine (benzoylecgonine), methamphetamine (methamphetamine), and heroin (6-acetylmorphine or morphine metabolites) [5]. Patients who were reported to have been prescribed any of these parent drugs or metabolites were excluded from analysis. Many patients were subject to multiple UDTs across time. Specimens were also excluded from analysis if they were collected within 30 days of a previous specimen. This filtering was performed to reduce the possibility that consecutive, closely timed, positive UDT results may derive from the same episode of drug use. A missing UDT result for a substance indicates it was not ordered by a practitioner.

We used the percentage of UDTs testing positive for a drug class as predictors in modeling. Illicit positivity rates were calculated by dividing the total number of positive results of a given drug by the total number of tests ordered for a given drug. Stimulant positivity was defined as having a positive test result for either methamphetamine or cocaine. Fentanyl co-positivity measures were derived by dividing the total number of samples positive for fentanyl and the co-positive drug by the number of samples positive for fentanyl and tested for the co-positive drug in question.

### Statistical Methods

#### I. Model

The model-building approach is summarized as follows: first, we developed a Bayesian hierarchical model that includes spatiotemporal random effects to investigate the appropriate underlying trends in space and time. Second, we included the selected variables in a chosen random effect model and compared prediction performance. For the random effect model selection, we explored different permutations of random effects to determine which model had the best prediction performance. The full model specification is expressed with:

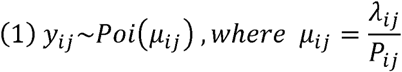

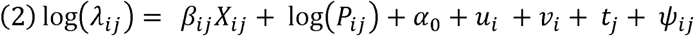

The equation (1) expresses *y_ij_* as the number of overdose deaths for county i=1…S, and time j=1…T, which follows a Poisson distribution with mean *μ_ij_*, which is the population-adjusted rate *λ_ij_* divided by county’s adult population (*P_ij_*) in each county i at time j. In the second hierarchy, the equation (2) models the population-adjusted rate (*λ_ij_*) with spatiotemporal random effects and fixed effects (*X_ij_*) (*β_ij_* are estimated parameters). The term *α*_0_ represents the baseline risk of overdose deaths, while *u_i_*+*v_i_* and *t_j_* are the random effects for space and time, respectively. The *u_i_*+*v_i_* random effect terms represent a Besag-York-Mollié (BYM) model [14], comprising a county-specific, independent and identically distributed(IID) random effect (unstructured) *v_i_* and a Besag (structured) random effect *u_i_*. The Besag model allows the spatial random effects of each county to correlate with their neighbors via an adjacency matrix [15]. If two counties are neighbors, the coordinate in the matrix takes on the value of 1; else the coordinate takes on the value of 0. The temporal term *t_j_* is modeled with a first-order random walk process (RW1) where previous time periods influence subsequent time periods *ψ_ij_* is the interaction effect between spatial and temporal effects and the four types of interaction effects were evaluated [16]. All random effects are organized in Table 1. The structure matrix of the interaction effects in the table can be obtained as the Kronecker product of the interacting main effects.

**Table 1.**
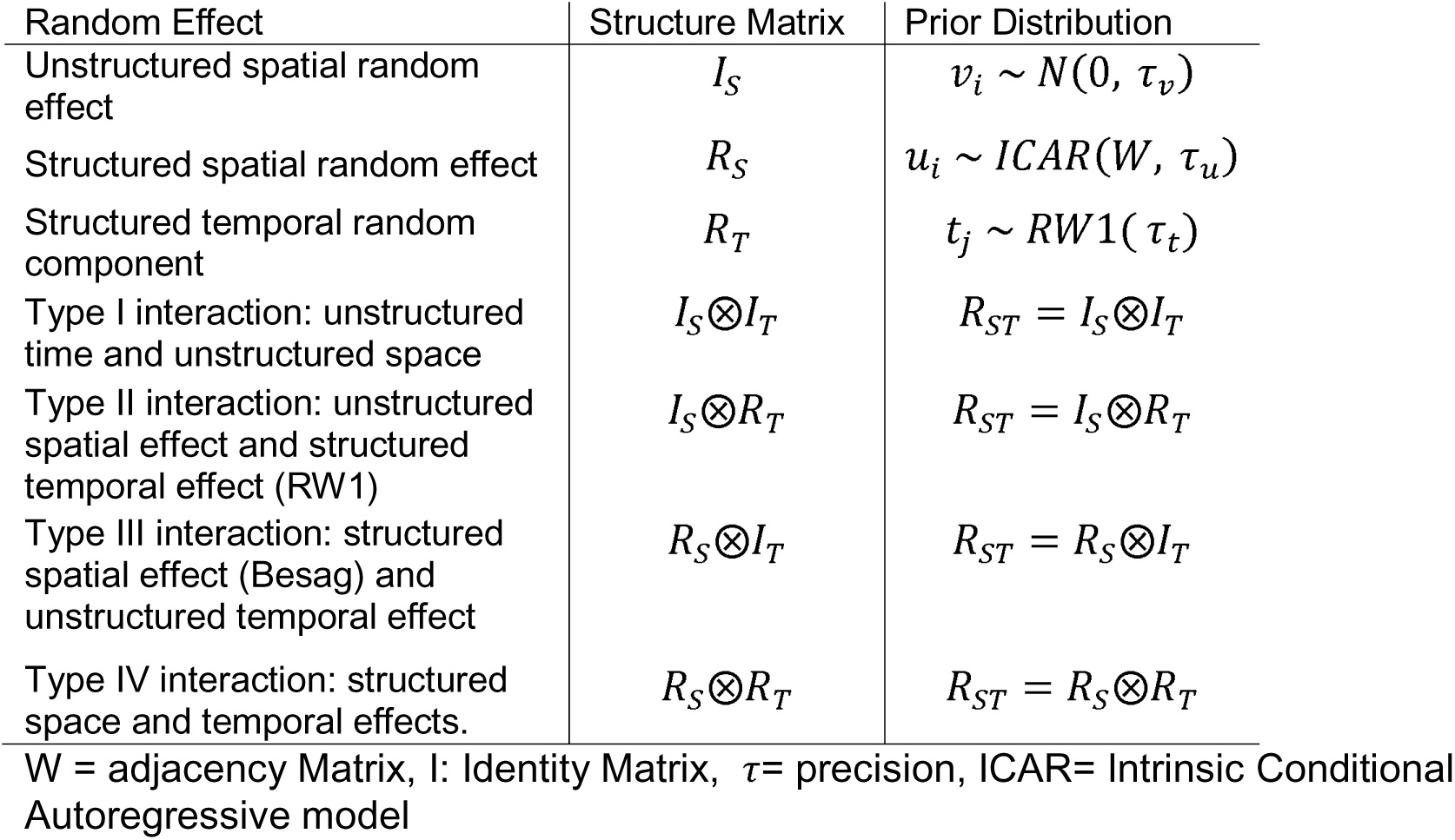
Specification of types of spatiotemporal random effects.

| Random Effect | Structure Matrix | Prior Distribution |
|---|---|---|
| Unstructured spatial random effect | *I<sub>S</sub>* | *v<sub>i</sub>* ~ *N*(0, *τ<sub>v</sub>*) |
| Structured spatial random effect | *R<sub>S</sub>* | *u<sub>i</sub>* ~ *ICAR*(*W*, *τ<sub>u</sub>*) |
| Structured temporal random component | *R<sub>T</sub>* | *t<sub>j</sub>* ~ *RW*1( *τ<sub>t</sub>*) |
| Type I interaction: unstructured time and unstructured space | *I<sub>S</sub>*⊗*I<sub>T</sub>* | *R<sub>ST</sub>* = *I<sub>S</sub>*⊗*I<sub>T</sub>* |
| Type II interaction: unstructured spatial effect and structured temporal effect (RW1) | *I<sub>S</sub>*⊗*R<sub>T</sub>* | *R<sub>ST</sub>* = *I<sub>S</sub>*⊗*R<sub>T</sub>* |
| Type III interaction: structured spatial effect (Besag) and unstructured temporal effect | *R<sub>S</sub>*⊗*I<sub>T</sub>* | *R<sub>ST</sub>* = *R<sub>S</sub>*⊗*I<sub>T</sub>* |
| Type IV interaction: structured space and temporal effects. | *R<sub>S</sub>*⊗*R<sub>T</sub>* | *R<sub>ST</sub>* = *R<sub>S</sub>*⊗*R<sub>T</sub>* |

W = adjacency Matrix, I: Identity Matrix,  *τ*= precision, ICAR= Intrinsic Conditional Autoregressive model

For the predictors, we selected several UDT variables for the model based on domain knowledge of opioid overdose death regarding drug usage trends, risk factors, and public health interventions. Fentanyl positivity was an obvious candidate due to its known associations with the third and fourth waves of the opioid crisis [17]. We also constructed a stimulant positivity measure as a positive result for either methamphetamine or cocaine. A co-positivity metric for fentanyl and stimulants – or stimulant positivity among fentanyl positive patients – was also utilized due to polysubstance use contributing to the fourth wave of the opioid crisis. We also used heroin and fentanyl-heroin co-positivity in the model. We noticed in the data that fentanyl-heroin co-positivity increased starting in 2023 as overdose death counts began to fall.

We used the overall SDI score, a metric composed of the other domains in the SDI. As SDI scores are percentiles, there is no guarantee that differences are consistent between percentiles (e.g. the difference between 10 and 20 is not necessarily the same as the difference between 20 and 30). We grouped the overall SDI score into four groups based on quantiles in Ohio (1-17, 18-39, 40-61, 62 or higher). We selected EMS events where naloxone was administered to serve as a proxy for potential opioid poisoning events, and we found that applying a natural logarithmic transformation to this metric yielded superior predictive performance and model fit compared to utilizing untransformed raw counts.

#### II. Model Implementation

We assessed our model’s performance using a rolling leave-future-out cross-validation (LFO-CV). This cross-validation method fits the model on a training set and uses future time periods as cross-validation to assess performance [18]. The prediction period was 2020 through 2024. We used a rolling window to keep the training set at a consistent size. For a visual example of the training-testing schema, refer to **Figure 1**.

**Figure 1.**
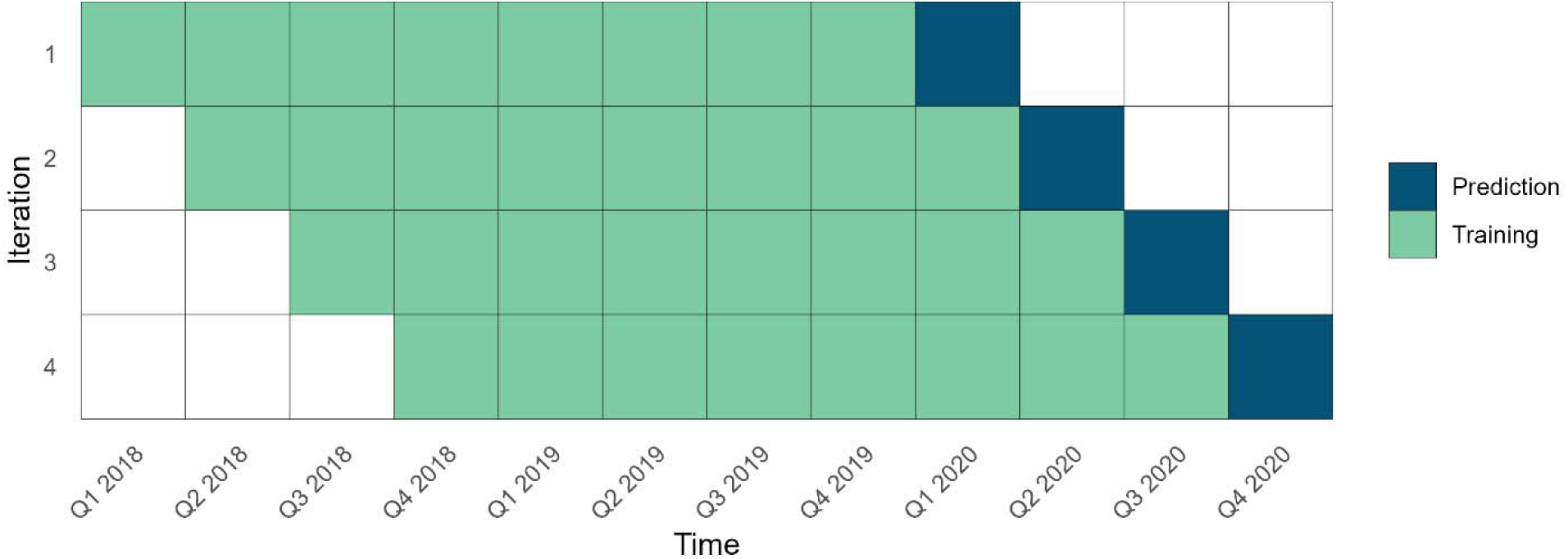
Model Validation Schema for 2020 using two years of training data.

Our rationale was that drug use trends evolve over time [17], and predictions may be less accurate with older data. We explored varying training windows ranging from four quarters to twelve quarters in length in four-quarter increments. Each iteration of the model was run on the specified number of quarters of training data per county. The model was then used to predict the next quarter of the training data. After prediction, the next model shifted the training window one quarter forward, incorporating a new quarter in the training set. Model performance was examined overall and grouped by year.

Model performance was assessed using several error terms: Mean Absolute Error (MAE), and Root Mean Square Error (RMSE). The formulas for these terms are as follows:

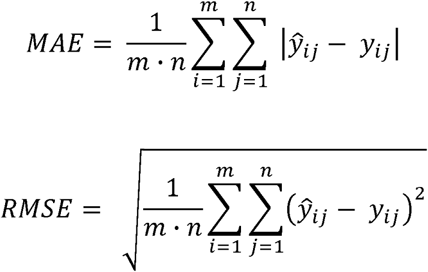

The term *y_ij_* is the observed number of overdoses for county i and time j, and *ŷ_ij_* is the predicted value of overdoses in all formulas. *m* is the number of counties in Ohio and *n* is the number of time periods being modeled. MAE is the mean absolute difference between observed and predicted values. RMSE is the square root of the mean squared difference between observed and predicted values; this metric weighs outlier predictions more heavily than MAE. Model performance results are presented both overall across all counties and stratified by county population size. Bayesian inference and model estimation for overdose mortality were performed using the Integrated Nested Laplace Approximation (INLA) approach[19]. INLA is a Bayesian inference method that approximates posterior distributions via Laplace Approximation as opposed to repeated simulations that Markov Chain Monte Carlo (MCMC) utilizes, which allows INLA to run markedly faster than MCMC. All analyses were conducted in R 4.5.3 [20], and the model implementation was conducted with the INLA package [21].

## Results

### Descriptive analysis of the data

Overdose death trends in the state of Ohio can be categorized into five distinct phases. Prior to 2016, there was a steady increase in opioid overdose deaths. The second phase is characterized by a surge in carfentanil occurring from 2016 to 2017. Overdose deaths surged and peaked in early 2017 before settling to their pre-surge levels at the beginning of 2018. From there, a gradual increase in overdose deaths continued until the onset of COVID-19. With the beginning of COVID-19, overdose deaths increased by approximately 30% at the state level from the first quarter (Q1) 2020 to the second quarter (Q2) 2020, and gradually fell over the course of subsequent years until the beginning of 2023. A sharp decline in overdose deaths started in mid-2023 and continued through the end of 2024. **Figure 2** displays state-level opioid overdose trends in Ohio. Maps displaying the average opioid death count per year and percent change from year to year can be found in Supplemental Figure 1.

**Figure 2.**
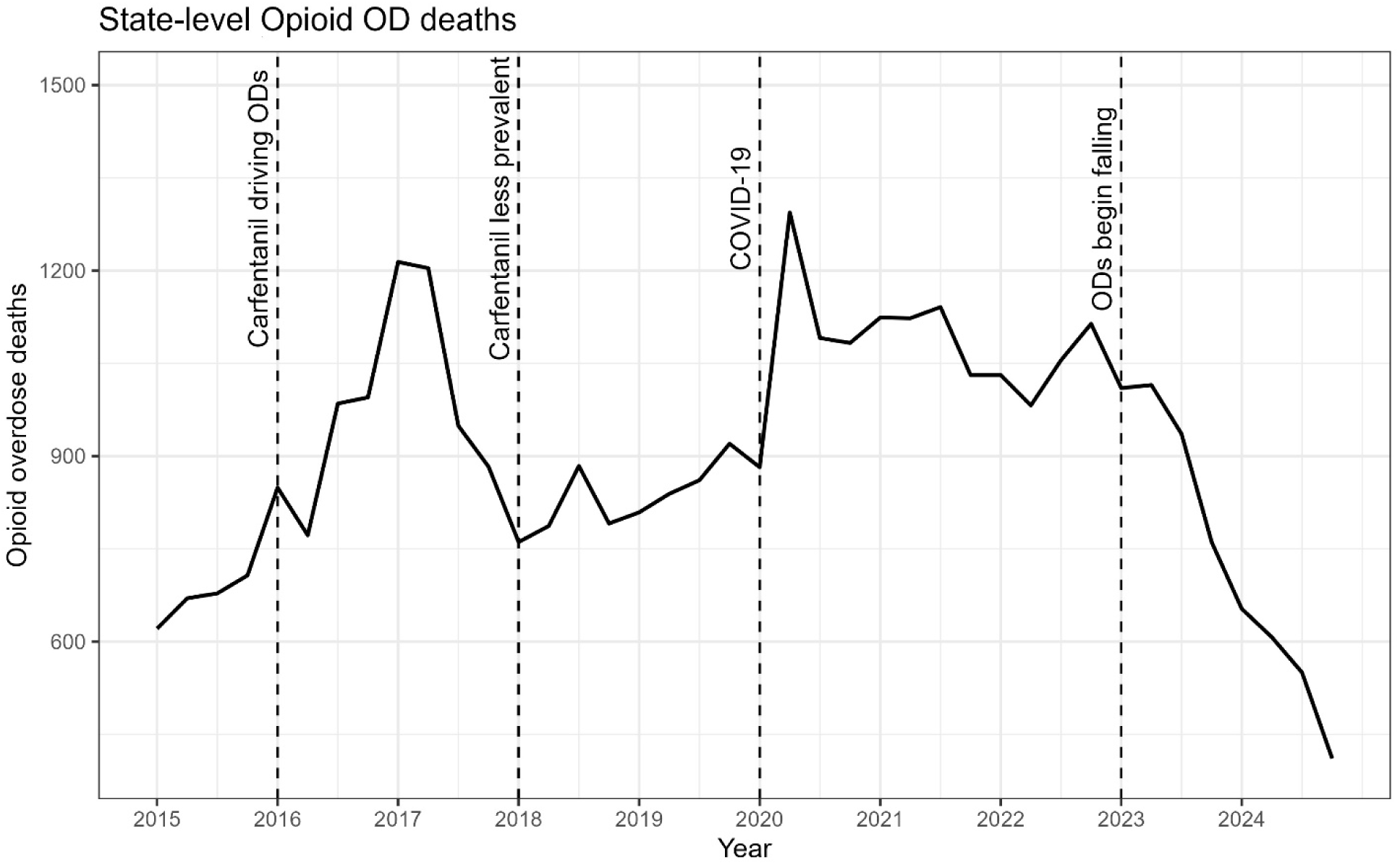
Opioid overdose deaths in Ohio, 2015-2024.

**Figure 3** displays the average UDT positivity rate (%) from 2015 to 2024 for select stimulants and fentanyl at the state level. Fentanyl positivity gradually increased over time until 2020, however, it doubled, from the Q1 2020 to the Q2 2020. It is noticeable that fentanyl positivity sharply decreased after that period and continuously decreased through 2024. Cocaine and methamphetamine experienced similar trends to a lesser degree. Regional differences exist in UDT positivity, with urban areas generally having a higher rate of cocaine use and rural regions having higher methamphetamine use. For more details on the change in UDT positivity over time, refer to Supplemental Figures 2-4. The number of samples of the UDT in each county weakly correlates with county population size.

**Figure 3.**
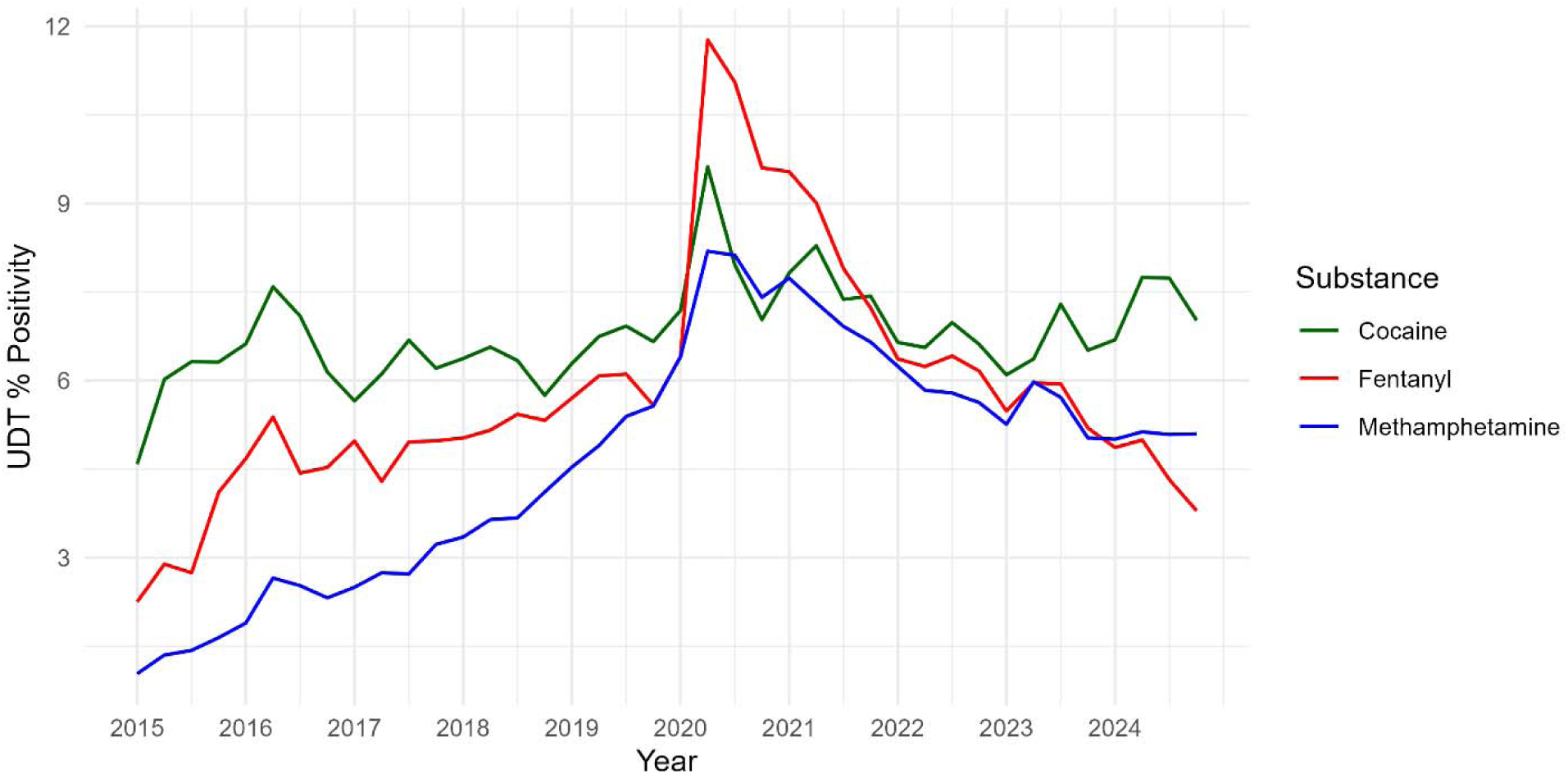
Fentanyl and select stimulant positivity aggregated to the state level

**Table 2** displays the county characteristics grouped by county size. Population, opioid overdose deaths, and UDT test volume per quarter are right-skewed metrics due to the influence of larger counties. The average county in Ohio has a population of approximately 103,000 adults, an SDI score of 40, over 10 opioid overdose deaths per quarter from 2015-2024 and 335 UDT tests per quarter. The median county in Ohio has a population of approximately 45,000, a similar SDI score, averages 3 opioid overdose deaths per quarter and 158 UDT tests per quarter. Population groupings were based on the mean (103,196) and median (44,884) adult population in counties. The county characteristics by geographic and urban/rural regions were provided in Supplemental Figure 5. In Ohio, northeast Ohio has the greatest population density, and counties adjacent to major metro centers tend to be more populous. Appalachian counties and major urban centers tend to have higher SDI scores, while western Ohio generally has lower (better) SDI scores.

**Table 2.**
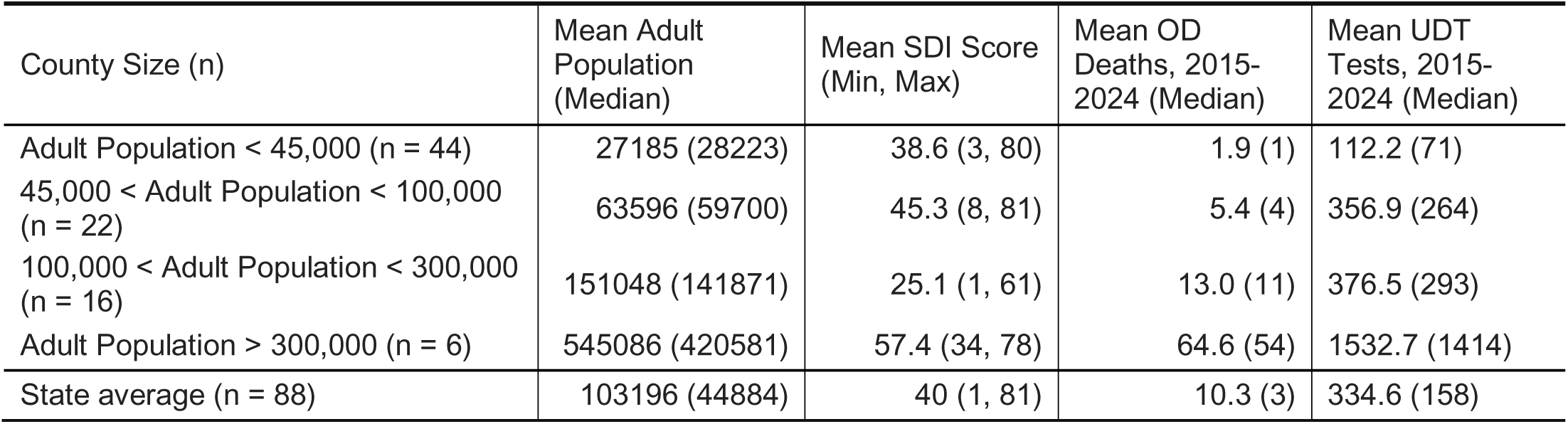
County characteristics grouped by county size.

| County Size (n) | Mean Adult Population (Median) | Mean SDI Score (Min, Max) | Mean OD Deaths, 2015-2024 (Median) | Mean UDT Tests, 2015-2024 (Median) |
|---|---|---|---|---|
| Adult Population < 45,000 (n = 44) | 27185 (28223) | 38.6 (3, 80) | 1.9 (1) | 112.2 (71) |
| 45,000 < Adult Population < 100,000 (n = 22) | 63596 (59700) | 45.3 (8, 81) | 5.4 (4) | 356.9 (264) |
| 100,000 < Adult Population < 300,000 (n = 16) | 151048 (141871) | 25.1 (1, 61) | 13.0 (11) | 376.5 (293) |
| Adult Population > 300,000 (n = 6) | 545086 (420581) | 57.4 (34, 78) | 64.6 (54) | 1532.7 (1414) |
| State average (n = 88) | 103196 (44884) | 40 (1, 81) | 10.3 (3) | 334.6 (158) |

### Prediction model development

Our initial phase was to determine which random effect specifications were the optimal baseline structure for the prediction model. We first used the data from 2020 through 2024 using only random effects with 8 quarters of training period and predicted the opioid overdose death of the next quarter. A 2-year training window was determined to be optimal (see Supplemental Table 1) in our sensitivity analysis. Supplemental Table 2 displays permutations of random effects and corresponding RMSE. For the temporal effect, RW1 has superior performance (overall RMSE = 8.20) and shows that the overdose death trend follows the trend of the previous time period.

Spatially, we found no meaningful improvement in prediction with more complex spatial structures such as BYM or BYM2 compared to the model with a single structured or unstructured effect (RMSE =4.89 to 4.90 across all models). With spatiotemporal main effects and four types of spatiotemporal interaction, the model with a Type IV interaction provided the best prediction performance among all models.

Following the selection of the optimal random effects structure, we integrated the fixed effects to construct the full predictive models and systematically compared their respective prediction performance. Table 3 displays the model performance stratified by county population size. MAE and RMSE are scale-dependent, and as such, their magnitude increases with the size of the outcome. For large metropolitan areas (population > 300,000), the comparison of models 1 and 2 shows that a large amount of variation remained unaccounted for after the baseline space and temporal random effects. Incorporating a Type IV spatiotemporal interaction term significantly mitigated this issue and reduced the MAE and RMSE. Models 3 to 6 investigated the effect of predictors on the prediction model. As the MAE and RMSE numbers showed, fentanyl positivity and naloxone events improved the prediction results, while other UDT data and SDI measures did not improve the prediction significantly. We finalized the prediction model by comparing Model 6 to the model without type IV random effects interaction (Model 7). Model 7 has marginally better performance than Model 6 and is more parsimonious. The change in overdose death information connected to the space-temporal interaction can be captured by the UDT and treatment information we provided.

**Table 3.**
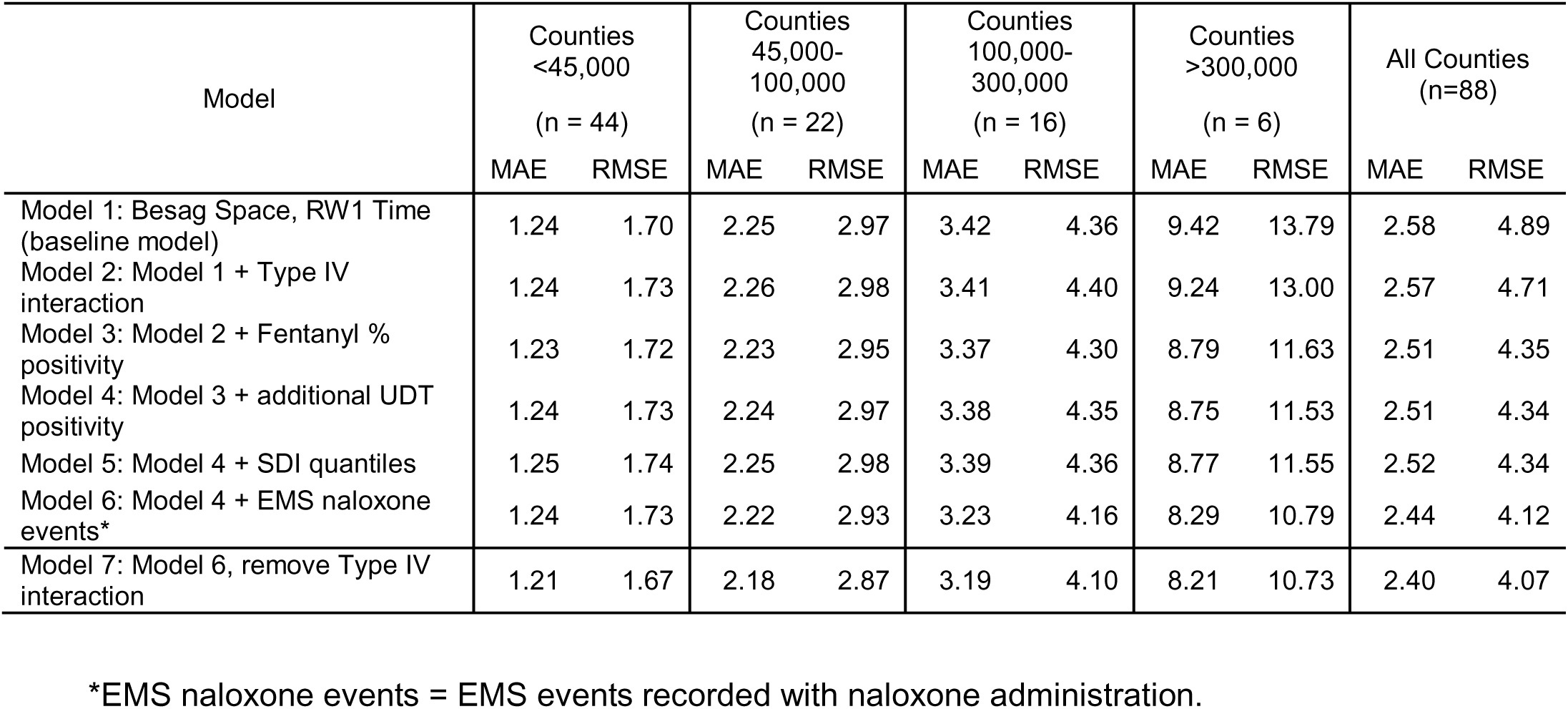
Model performance stratified by adult population and model specification, 2020-2024. Successive models build on previous models, notated by the model number.

| Model | Counties <45,000<br>(n = 44) |  | Counties 45,000-100,000<br>(n = 22) |  | Counties 100,000-300,000<br>(n = 16) |  | Counties >300,000<br>(n = 6) |  | All Counties (n=88) |  |
|---|---|---|---|---|---|---|---|---|---|---|
|  | MAE | RMSE | MAE | RMSE | MAE | RMSE | MAE | RMSE | MAE | RMSE |
| Model 1: Besag Space, RW1 Time (baseline model) | 1.24 | 1.70 | 2.25 | 2.97 | 3.42 | 4.36 | 9.42 | 13.79 | 2.58 | 4.89 |
| Model 2: Model 1 + Type IV interaction | 1.24 | 1.73 | 2.26 | 2.98 | 3.41 | 4.40 | 9.24 | 13.00 | 2.57 | 4.71 |
| Model 3: Model 2 + Fentanyl % positivity | 1.23 | 1.72 | 2.23 | 2.95 | 3.37 | 4.30 | 8.79 | 11.63 | 2.51 | 4.35 |
| Model 4: Model 3 + additional UDT positivity | 1.24 | 1.73 | 2.24 | 2.97 | 3.38 | 4.35 | 8.75 | 11.53 | 2.51 | 4.34 |
| Model 5: Model 4 + SDI quantiles | 1.25 | 1.74 | 2.25 | 2.98 | 3.39 | 4.36 | 8.77 | 11.55 | 2.52 | 4.34 |
| Model 6: Model 4 + EMS naloxone events* | 1.24 | 1.73 | 2.22 | 2.93 | 3.23 | 4.16 | 8.29 | 10.79 | 2.44 | 4.12 |
| Model 7: Model 6, remove Type IV interaction | 1.21 | 1.67 | 2.18 | 2.87 | 3.19 | 4.10 | 8.21 | 10.73 | 2.40 | 4.07 |

\*EMS naloxone events = EMS events recorded with naloxone administration.

### Prediction Performance by Year

From 2020 to 2024, the trend of opioid overdose deaths showed very different patterns each year. The year 2020 shows a large increase, and 2021-2023 show a gradual decrease, and a sudden decrease starting in 2023. We conducted a detailed investigation of each year to show how our model performs in different overdose death trends.

Table 4 shows model performance presented by year and model specification. RMSE was selected to assess model performance because its squared-error formulation assigns greater weight to larger prediction errors, making it particularly useful for identifying poor predictive performance. As with the overall result, Model 7 shows the best performance for most of the years. For the years with high volatility (2020, 2023, and 2024), models incorporating fixed predictors (Models 3-7) show better performance than Models 1 and 2. However, in the years with gradual changes (2021 and 2022), we can observe that the effect of fixed predictors was diminished. It is also notable that when we observed the large increasing trend (2020), fentanyl positivity and the EMS naloxone variables had a substantial effect on prediction, where RMSE decreased by 2 from Model 1 (an almost 27 percent decrease) and by 1.37 from Model 2 (almost 20 percent decrease). Other UDT positivity (heroin, other stimulants, and their co-positivity with fentanyl) was redundant in 2020 (Model 4), but when we observed the large decreasing trend in 2023 and 2024, other UDT positivity data contributed modestly to the prediction after accounting for fentanyl positivity.

**Table 4.** Model performance stratified by year and model specification.

| Model | RMSE<br>2020 | RMSE<br>2021 | RMSE<br>2022 | RMSE<br>2023 | RMSE<br>2024 | Overall<br>RMSE |
|---|---|---|---|---|---|---|
| Model 1: Besag Space, RW1 Time<br>(baseline model) | 7.62 | 3.75 | 3.79 | 4.43 | 3.69 | 4.89 |
| Model 2: Model 1 + Type IV interaction | 6.94 | 3.82 | 3.78 | 4.44 | 3.76 | 4.71 |
| Model 3: Model 2 + Fentanyl % positivity | 5.57 | 3.81 | 3.89 | 4.46 | 3.74 | 4.35 |
| Model 4: Model 3 + additional UDT positivity | 5.62 | 3.78 | 3.97 | 4.35 | 3.67 | 4.34 |
| Model 5: Model 4 + SDI quantiles | 5.63 | 3.78 | 3.97 | 4.37 | 3.67 | 4.34 |
| Model 6: Model 4 + EMS naloxone events | 5.28 | 3.66 | 3.96 | 4.14 | 3.29 | 4.12 |
| Model 7: Model 6, remove Type IV interaction | 5.24 | 3.68 | 4.01 | 4.06 | 3.06 | 4.07 |

One of the desirable features of a prediction model is to accurately predict the beginning of the outbreak (i.e., a sudden increase in cases). We examine the early detection ability of our model with the data from 2020. The second quarter of 2020 recorded a large increase in opioid deaths corresponding to the beginning of the COVID-19 pandemic (Figure 2). Table 5 shows model performance stratified by the quarters of 2020. Our primary focus is on 2020 Q2, evaluating how well each model predicted the sharp increase observed at the onset of the COVID-19 pandemic. For Q1, the spatiotemporal random effects alone (Models 1 and 2) yielded a comparable fit to the models incorporating fixed effects. However, during Q2, the baseline models (Models 1 and 2) exhibited a dramatic increase in RMSE, demonstrating that purely random-effects formulations are insufficient to capture sudden, volatile shifts in overdose trends. The integration of fentanyl UDT positivity substantially enhanced predictive accuracy, most notably reducing the RMSE in Q2 by 35% relative to Model 2. Furthermore, incorporating EMS naloxone administration events provided additional incremental improvements in model performance.

**Table 5.** Model performance in 2020.

| Model | RMSE<br>Q1 | RMSE<br>Q2 | RMSE<br>Q3 | RMSE<br>Q4 | Overall<br>RMSE |
|---|---|---|---|---|---|
| Model 1: Besag Space, RW1 Time(baseline model) | 4.57 | 11.76 | 6.72 | 5.29 | 7.62 |
| Model 2: Model 1 + Type IV interaction | 4.57 | 10.72 | 6.04 | 4.48 | 6.94 |
| Model 3: Model 2 + Fentanyl % positivity | 4.73 | 6.99 | 5.99 | 4.15 | 5.57 |
| Model 4: Model 3 + additional UDT positivity | 4.90 | 6.87 | 5.90 | 4.51 | 5.62 |
| Model 5: Model 4 + SDI quantiles | 4.98 | 6.78 | 5.95 | 4.52 | 5.63 |
| Model 6: Model 4 + EMS naloxone events | 4.56 | 6.19 | 5.64 | 4.52 | 5.28 |
| Model 7: Model 6, remove Type IV interaction | 3.68 | 6.24 | 5.90 | 4.74 | 5.24 |

Gray columns indicate time periods after COVID-19 began.

### Prediction results in selected counties

In this section, we investigated the predictive model performance for each county. Table 6 and Figure 4 display model predictions for Williams, Erie, Clermont, and Franklin counties with the best-performing model (Model 7). For each population group, we selected a representative county whose adult population was nearest to the median adult population within that group (see Table 2). Williams represents counties with populations smaller than the median population (45,000) of the state. Erie represents counties with a population of more than the median and less than the mean population (103,196). Clermont represents counties that are larger than the state’s mean population but are not major metro areas. We defined major metro areas as the counties containing Columbus, Cleveland, Cincinnati, Akron, Dayton, and Toledo. Franklin County is representative of this group.

**Figure 4.**
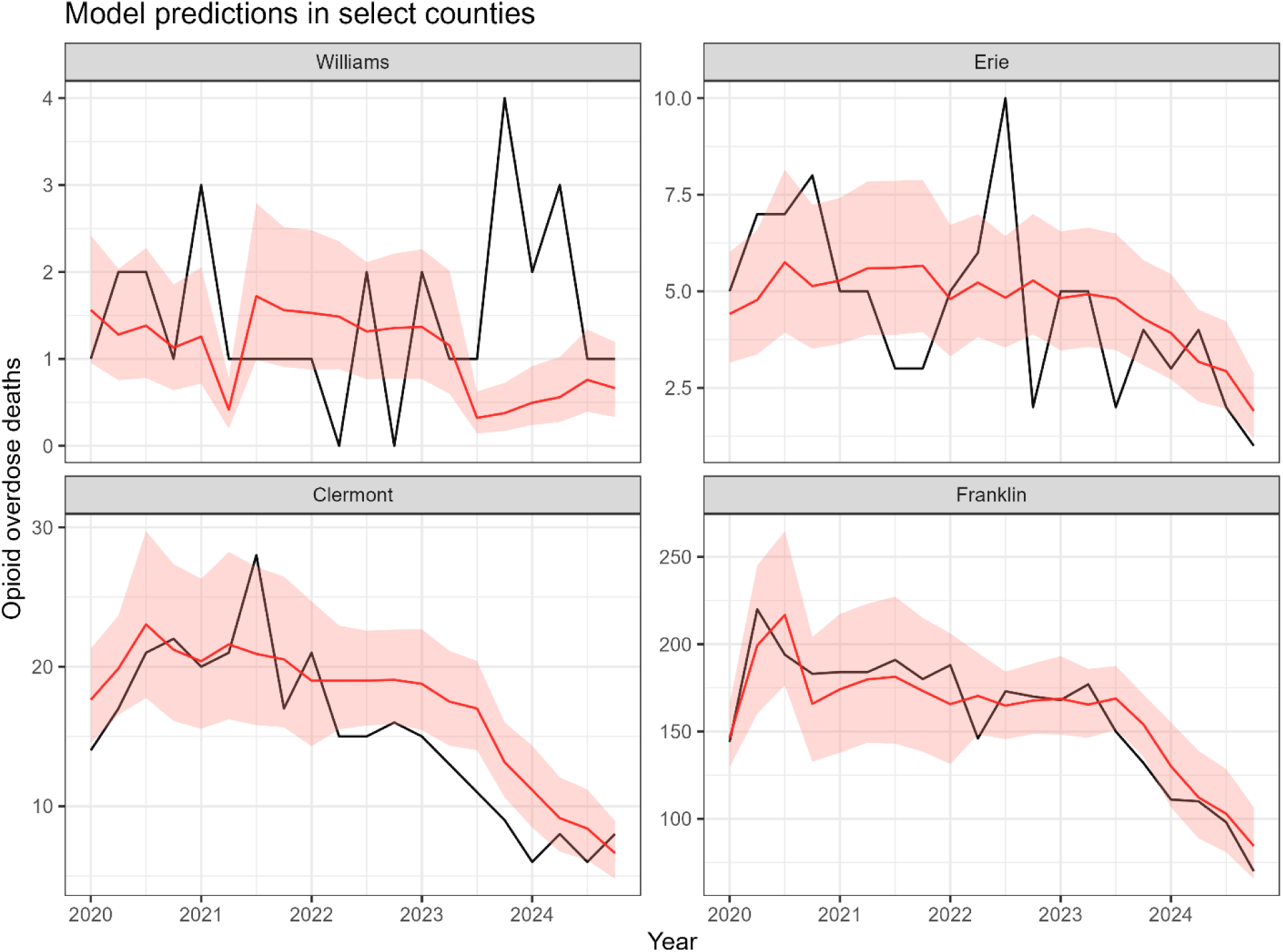
Model predictions for select counties. Black lines are the observed opioid overdose deaths, red lines are predicted deaths, and the salmon-shaded bands are the 95% Credible Intervals.

**Table 6.** Model performance in select counties, 2020-2024.

| County | Average adult population | Mean OD count per quarter, 2020-2024 | Mean OD rate per 100k adults | SDI | MAE | RMSE |
|---|---|---|---|---|---|---|
| Williams | 28603 | 1.5 | 5.2 | 17 | 0.99 | 1.32 |
| Erie | 59842 | 4.6 | 7.7 | 27 | 1.44 | 1.97 |
| Clermont | 157646 | 15.2 | 9.6 | 13 | 3.06 | 3.54 |
| Franklin | 984709 | 158.7 | 16.1 | 66 | 12.13 | 14.73 |

Our results demonstrate that model performance varies systematically with county population size and caseload. Franklin and Clermont counties have stronger and more stable prediction performance compared to the other two small counties. While their MAE values are larger in absolute terms, these errors were proportionally small relative to the high volume of observed overdose deaths, with many predictions falling securely within the 95% Bayesian credible intervals. However, Williams and Erie counties exhibit greater prediction volatility, resulting in MAEs that are proportionally larger relative to their lower observed mortality counts. They have an average overdose death of less than 5, and this limited data causes wider posterior credible intervals with greater parameter uncertainty. Crucially, however, our proposed model (model 7) consistently outperformed the spatiotemporal random-component baseline within these smaller, highly volatile counties (Table 3).

## Discussion

In this study, we investigated the Bayesian spatiotemporal modeling approach to predict opioid overdose deaths in Ohio from 2020 to 2024. We utilized a rolling 2-year training window to predict overdose deaths in the next quarter, a strategy specifically designed to provide timely estimates despite the inherent 2-to-4-month lag in official mortality reporting. Our best-performing model combined a structured spatial random effect (Besag) and a correlated temporal random effect (RW1) as the baseline structure, integrated with fixed predictors for UDT metrics and EMS naloxone events.

We conducted an extensive investigation of the prediction model with random effects and predictors. While a Type IV spatiotemporal interaction initially enhanced model fit, subsequent evaluation of the model showed that the model with main spatiotemporal effects and UDT and EMS data achieved similar or better performance without interaction. Type IV interaction (interaction between structured spatial and structured temporal effects) is related to localized, time-specific deviations from the main spatial and temporal trends. MAE and RMSE results indicated that we systematically account for the measurable drivers of these localized sudden changes by using the UDT data (especially fentanyl) and EMS naloxone administration data. Fentanyl is a known primary contributor to overdose deaths, and our fentanyl UDT data reveal significant spatiotemporal heterogeneity in its prevalence across Ohio. The predictive power of the UDT data was more pronounced in model performance in the year 2020, with the most substantial improvements in model predictive performance during this period. Opioid-related overdose deaths surged in 2020, with deaths increasing most drastically from Q1 2020 to Q2 2020. The analysis of the year 2020 demonstrates that the utilization of the UDT – particularly fentanyl - in our prediction model enables the model to effectively accommodate the sharp increase in Q2 and gives significantly enhanced predictive performance. Between 2023 and 2024, opioid overdose deaths showed a significant downward trend. UDT data did not provide much improvement to model performance, but EMS naloxone administration data provided substantially improved prediction results when the large shift occurred. Therefore, leveraging these rapid-reporting surveillance sources effectively captures acute, localized fluctuations while significantly enhancing overall model forecasting accuracy.

We also observed that prediction performance exhibited significant heterogeneity, varying systematically according to the population size of each county. As discussed in the model performance in select counties, the largest counties have the best model performance. Crucially, this aligns with the observation that the most critical surges and sharp increases in overdose deaths were predominantly localized within major metropolitan areas. For example, during Q2 2020, opioid overdose deaths increased by over 30% in Franklin County. Consequently, the model is structurally optimized to capture these high-volume, structural trends, resulting in relatively minimized relative error margins in the specific counties facing the highest public health burden. In addition, the high noise-to-signal ratio in low-population areas makes it challenging to forecast small-scale shifts, potentially influencing the specification of the spatial random effects. A substantial proportion of regions exhibit low overdose mortality counts; specifically, 44 out of 88 counties averaged 1.9 overdose deaths per quarter during 2015 to 2024 (Table 2). Given these low event counts, raw empirical estimates in these counties are highly unstable. To mitigate this problem, previous overdose death research used a framework where small Ohio counties were classified by their likelihood of crossing an outcome threshold [6].

## Conclusion

This study developed a robust Bayesian spatiotemporal forecasting framework to predict county-level, quarterly opioid overdose mortality in Ohio from 2020 to 2024. Our predicted results bridge the critical 2-to-4-month reporting lag inherent to official mortality databases. The optimal model configuration integrated spatiotemporal random effects with rapid-reporting surveillance data (UDT and EMS naloxone data). These dynamic data streams contributed significantly to overall predictive accuracy, demonstrating their greatest utility during sudden and substantial shifts in opioid overdose mortality. UDT data represents a promising surveillance resource for the development of an early warning system. To our knowledge, these data have not previously been subjected to rigorous analysis with advanced statistical methods at a finer geographic scale than the regional/state-level data, aside from our previous machine learning prediction model paper [6]. Therefore, we anticipate that future studies will leverage these datasets not only to optimize the prediction of localized opioid overdose death trends, but also to investigate evolving drug overdose risk factors, substance use, and behavioral patterns.

One limitation of our study is that the availability of the UDT data is not consistent across the Ohio counties. While we have samples available across the state, the volume of test samples loosely correlates with population size. Small counties generally have fewer UDT tests available, so we lack resolution on drug use trends in those areas. Future studies can incorporate the modeling approach to mitigate the effect of a sparse sample of UDT, similar to the small area estimation approaches to stabilize these local indicators. Another limitation is regarding the geographical region and temporal trend. The variables we selected, especially for the near-real-time data, were chosen based on the State of Ohio. Variable selection and relevant prediction models would likely be different in other states, as underlying drug use trends could result in different associations between UDT positivity and opioid overdose deaths. Our prediction period was limited to 2020 to 2024, and the UDT variables we selected were chosen based on known overdose trends in Ohio, so drug overdose trends and socioeconomic situation are characteristics of this period. We anticipate that future studies can adapt these rapid reporting surveillance data for rigorous predictive models in alternative geographic regions and time periods.

The primary utility of our prediction model lies in its capacity to serve as the foundation for an early warning system during acute surges in opioid overdose mortality. Ultimately, this model can equip public health practitioners with the predictive precision required to allocate resources for timely and targeted interventions.

## Data Availability

Sociodemographic and Census metrics are publicly available and can be accessed directly through the U.S. Census Bureau website (data.census.gov) and the Robert Graham Center Social Deprivation Index (SDI) repository (graham-center.org).The mortality data are proprietary to the Ohio Department of Health (ODH) and are
subject to legal and ethical restrictions to protect patient privacy. Qualified researchers may apply for access to these data directly through the ODH Institutional Review Board or data request portal at https://data.ohio.gov/wps/portal/gov/data/. Aggregated forms
of the mortality data are available through the Ohio Integrated Behavioral Health Dashboard.
(https://data.ohio.gov/wps/portal/gov/data/view/ohio-ibhd).
EMS-related data are available through the Ohio Integrated Ohio Behavioral Health Dashboard. The Urine Drug Test (UDT) data are proprietary to Millennium Health LLC and were obtained under a specific data-sharing agreement for this research. An aggregated
form of drug positivity data can be available upon request. Other researchers may request access to these data independently by contacting Millennium Health LLC at https://www.millenniumhealth.com/.

