## Supplemental Information for "Predicting county-level opioid overdose mortality in Ohio through multimodal data: A spatiotemporal study using toxicology and EMS indicators, 2020-2024"

**Supplemental Figure 1**. Opioid overdose deaths from 2015 to 2024 and % change in overdose deaths year after year.

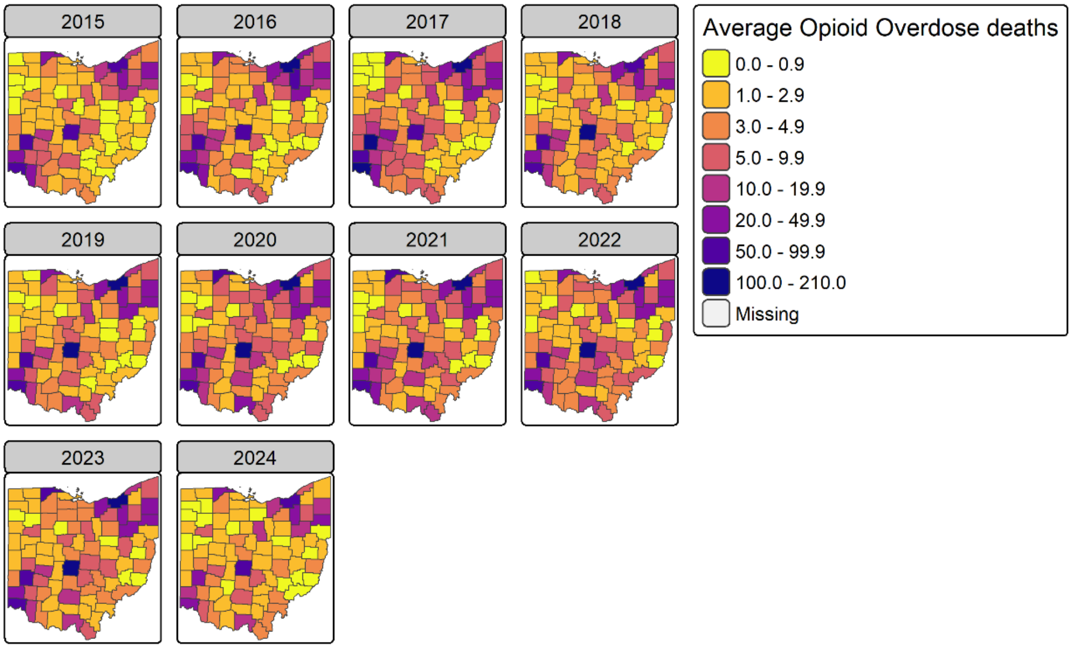


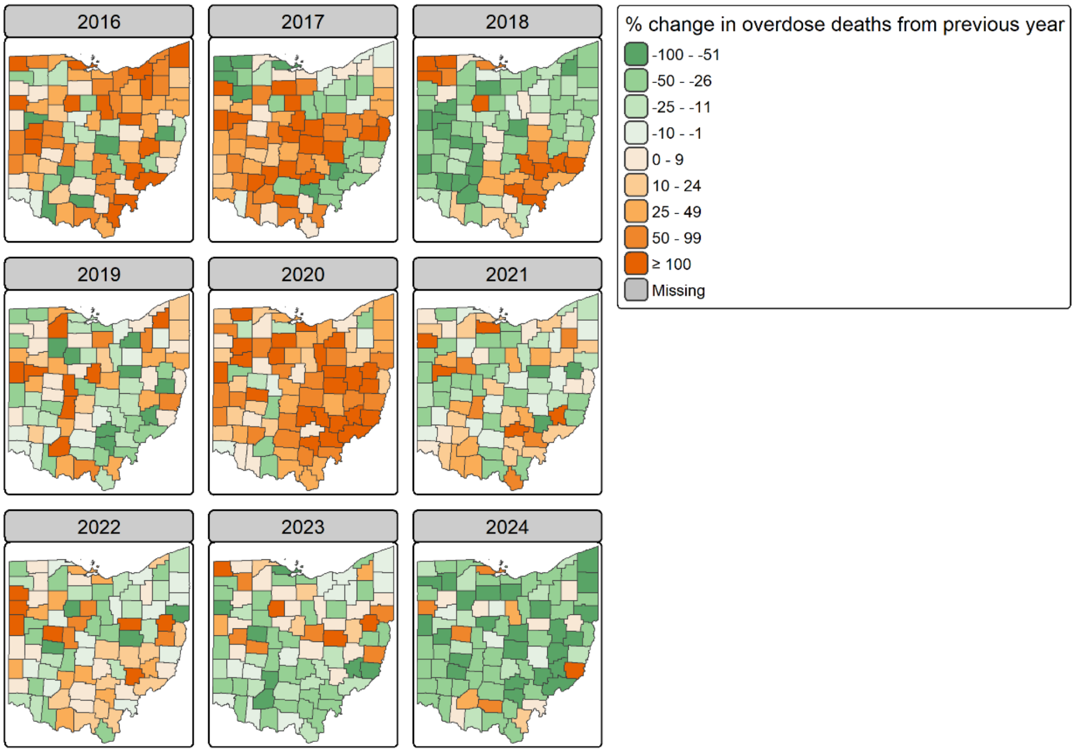

**Supplemental Figure 2**. Fentanyl positivity over time, 2015-2024.

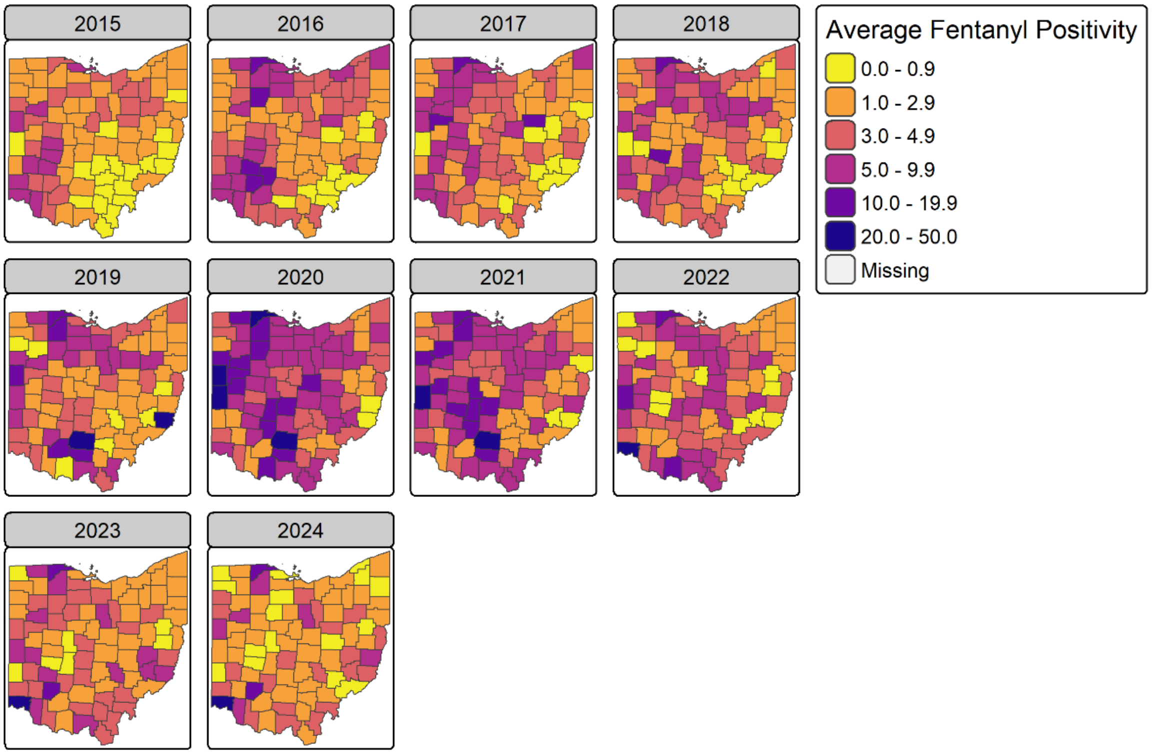

**Supplemental Figure 3**. Average Methamphetamine Positivity, 2015-2024.

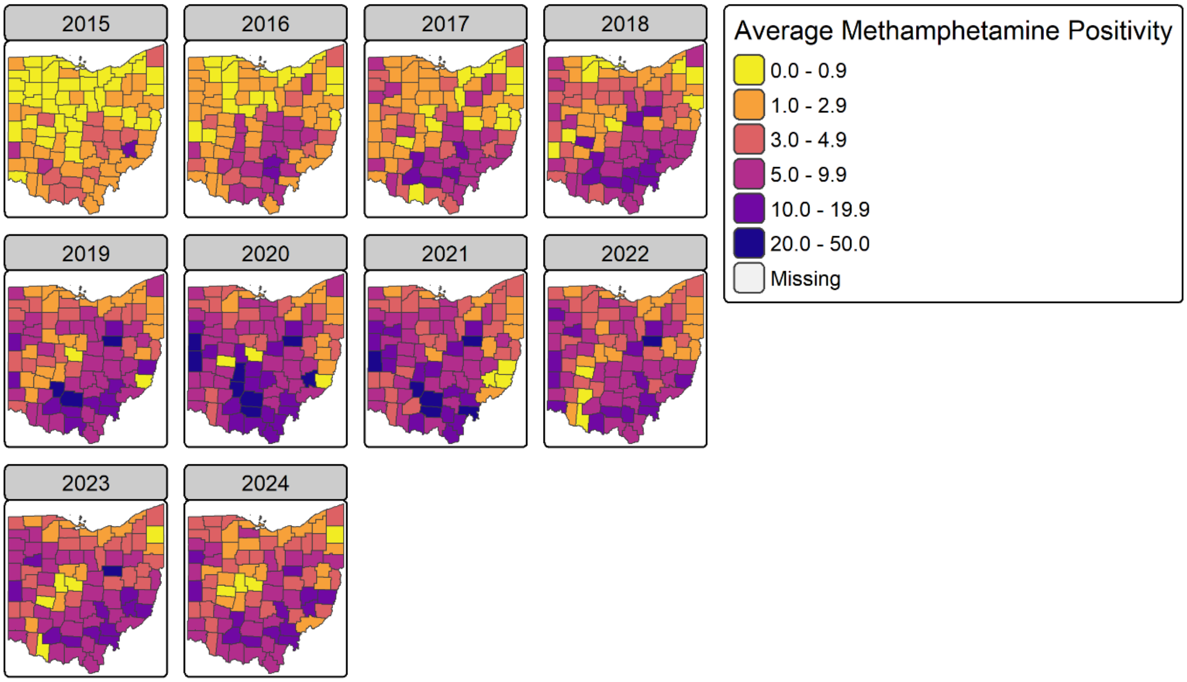

**Supplemental Figure 4.** Average Cocaine Positivity, 2015-2024.

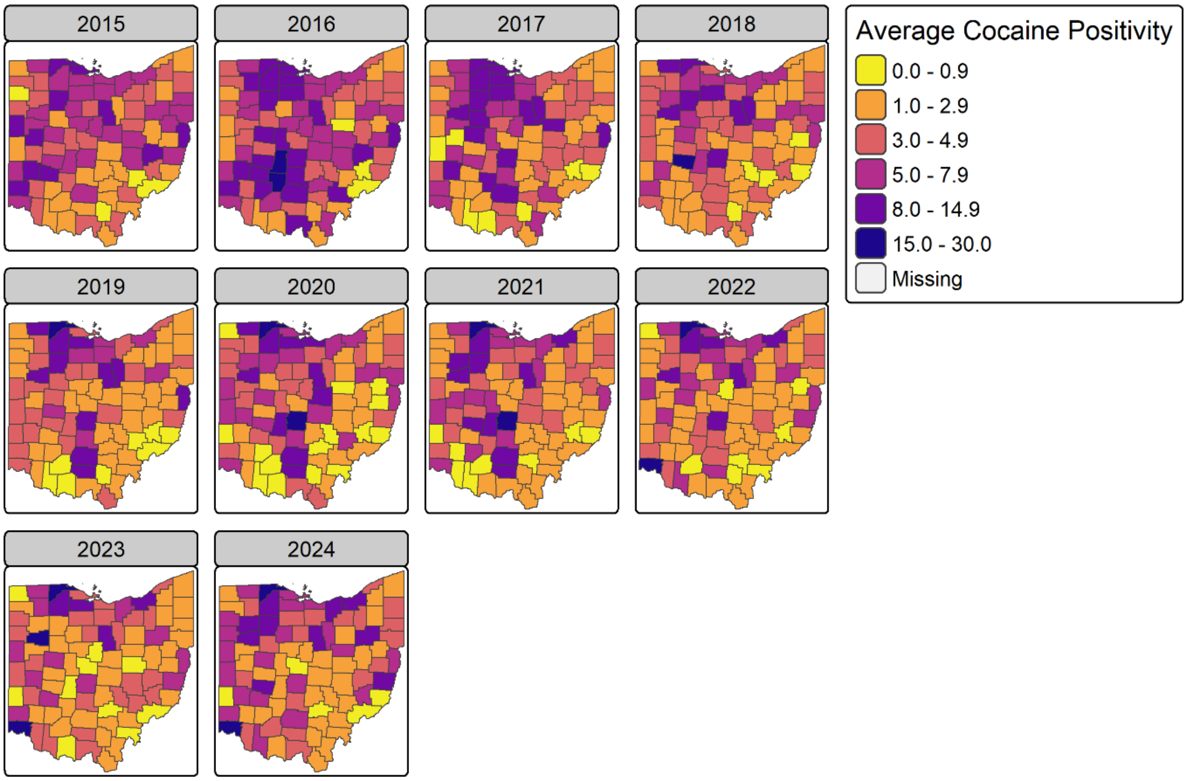

**Supplemental Figure 5**. Average adult population and overall SDI score by county. SDI scores are grouped by quantiles.

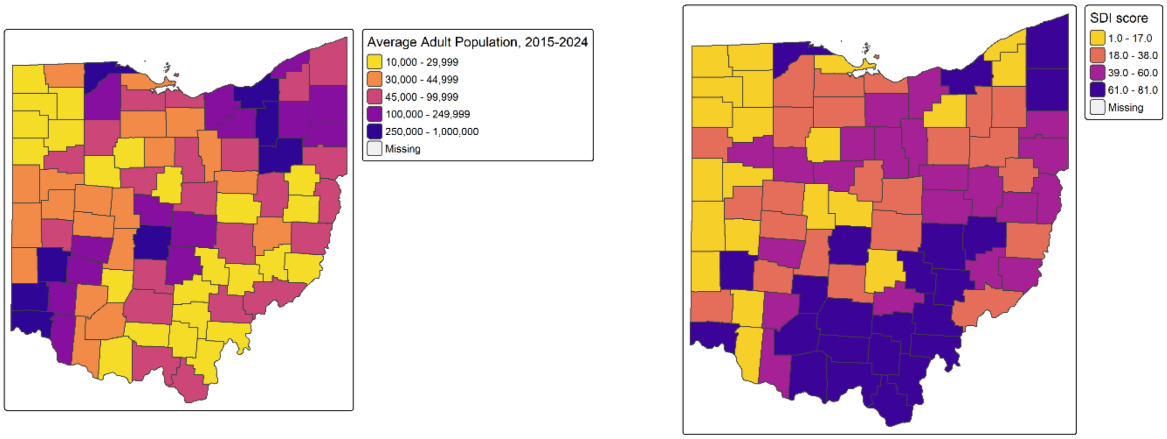

**Supplemental Table 1**. Model performance by training set length, averaged over 2020-2024.

| Training Length | MAE | RMSE |
|---|---|---|
| 4 quarters | 2.48 | 4.19 |
| 8 quarters | 2.42 | 4.09 |
| 12 quarters | 2.42 | 4.18 |

**Supplemental Table 2**. Random effect models predicting 2020-2024.

| Model specification, 2020-2024 | RMSE |
|---|---|
| Time IID | 9.05 |
| Time RW1 | 8.20 |
| Time AR1 | 8.47 |
| County IID | 6.60 |
| County Besag | 6.60 |
| County BYM | 6.61 |
| County BYM2 | 6.60 |
| Time RW1, County IID | 4.89 |
| Time RW1, County Besag | 4.89 |
| Time RW1, County BYM | 4.90 |
| Time RW1, County BYM2 | 4.89 |
| Time RW1, County IID, Type I Interaction | 4.93 |
| Time RW1, County IID, Type II Interaction | 4.77 |
| Time RW1, County Besag, Type III Interaction | 4.98 |
| Time RW1, County Besag, Type IV Interaction | 4.71 |

All models have a Poisson distribution and an offset of the log of county population.
